# Impact of the first nirsevimab immunization campaign on RSV-related emergency department visits and hospitalizations in Québec, Canada

**DOI:** 10.1101/2025.10.20.25338243

**Authors:** Amandine Bemmo, Élise Fortin, Patrick Fortin, Radhouene Doggui, Charles-Antoine Guay, Rachid Amini, Sara Carazo, Jesse Papenburg, Rodica Gilca

**Author notes:** Corresponding author: Élise Fortin, Direction des risques biologiques, Institut national de santé publique du Québec 945, avenue Wolfe, Québec, Québec. Co-first authors (equally contributed) (Amandine Bemmo, Élise Fortin). **Conflicts of interests**: RG reports funding from the *Ministère de la Santé et des Services sociaux du Québec* paid to her institution for this study. JP reports grants from MedImmune and Merck to his institution, and personal fees from Enanta, all outside the submitted work. EF reports funding from the *Ministère de la Santé et des Services sociaux du Québec* paid to her institution, but nor pertaining to the current study. SC reports funding from Public Health Agency of Canada for COVID-19 studies paid to her institution, but nor pertaining to the current study. Other authors have no conflicts of interests to declare. **Availability of data and code:** data was obtained through governmental mandate and cannot be shared by the authors. Any requests should be addressed to the *Ministère de la Santé et des Services sociaux du Québec.* Code can be shared upon request by the authors. **Acknowledgements:** we would like to thank the HospiVir coordinating team as well as the local teams from participating hospitals. We are also grateful to the INSPQ’s *Infocentre de santé publique* team, who provided data extractions for administrative databases.

## Abstract

**Objectives:** During the 2024-2025 respiratory syncytial virus (RSV) season, a universal nirsevimab infant immunization program was implemented in the province of Québec. We evaluated its population-level impact on RSV-related emergency department (ED) visits and hospitalizations in infants, using both active surveillance and administrative health data.

**Methods:** Study population included all Québec children <18 years old. Incidence rates of RSV-confirmed hospitalizations (6 hospital-based active surveillance), RSV-associated hospitalizations and ED visits linked to a positive RSV test, and acute bronchiolitis ED visits (administrative databases) were measured for nirsevimab-eligible (0-5 months old) and non-eligible (6-11 months and 1-17 years old) age groups during the 2024-2025 season and compared to the previous seasons (varying lookback periods). Using difference-in-differences and observed versus expected approaches, pre-/post-intervention variations in 0-5-month-olds were adjusted for any time trends extending to non-targeted age groups.

**Results:** Decreases of 59% (95% CI: 49–70) and 66% (95% CI: 58–71) were observed in RSV-confirmed and RSV-associated hospitalizations, respectively. Acute bronchiolitis ED visits decreased by 35% (95% CI: 28–41), and RSV-associated ED visits, by 60% (95% CI: 56–65). This impact became noticeable approximately one week after expanding the campaign to all eligible infants. ED visits and hospitalizations were less frequent than expected (i.e. without nirsevimab) in 0-5-month-olds, across all indicators. Between 323 and 746 hospitalizations were possibly prevented.

**Conclusion:** The 2024-2025 nirsevimab campaign was associated with a two-thirds reduction in RSV-related hospitalizations and RSV-associated ED visits in Québec infants. ED visits for acute bronchiolitis were also reduced by a third.

## Introduction

Respiratory syncytial virus (RSV) infections cause substantial morbidity in young children, especially infants. The global estimated burden among children 0 to 5 years old was of 3.6 millions of RSV-associated hospitalizations and 100,000 deaths in 2019.^1^ Between 40% and 45% of these involved infants younger than 6 months.^1^ In the Province of Québec, during the 2022-2023 season (before the RSV immunization program), RSV was the most frequent cause of pediatric hospitalization for an acute respiratory infection (ARI; 39% of ARI hospitalizations).^2^

Nirsevimab, a monoclonal antibody conferring passive immunity against RSV infections, was authorized in Canada for children under 2 years old in April 2023^3^. Unlike palivizumab, which required monthly injections and was reserved for high-risk infants, nirsevimab may provide broader protection to all infants, with a single dose per RSV season. Starting in Fall 2024, nirsevimab was publicly funded and recommended for all infants born during 2024-2025 season or younger than 6 months at the beginning of the season.^4^ It was also offered to infants 6-18 months old with certain conditions, as well as to those living in remote communities. Québec’s immunization campaign began on October 1^st^, 2024, reaching around 70% coverage by the end of the campaign in infants less than 6 months old.^5^ Nirsevimab’s effectiveness in preventing RSV severe outcomes has been largely documented.^6^ However, few studies have measured the real-life populational impact on infant RSV-related hospitalizations, adjusting with comparison groups for other temporal variations such as year-to-year variations in virus virulence and RSV attack rates.^7–13^ None have described the impact on emergency department (ED) visits. These studies report a wide range of reductions in hospitalizations, depending in large part on immunization coverage. Implementation of the immunization campaign in Québec might differ from what was done elsewhere, potentially affecting its impact.

We estimated the populational impact of the nirsevimab immunization campaign in Québec on the frequency of RSV-related ED visits and hospitalizations among infants <6 months old, using both administrative health data and active surveillance data. The comparison of results obtained using different data sources and case definitions will inform on the potential consequences of methodological choices when evaluating an immunization campaign’s impact.

## Methods

### Study design and population

This quasi-experimental study included children <18 years old from the Province of Québec, except for data from the HospiVir active surveillance network which included only children seen at participating hospitals. Impact was measured on nirsevimab-eligible infants <6 months old (according to age at ED visit or admission or ED visit) and compared to children 1-17 years old. Children 6-11 months old were included in sensitivity analyses, as immunization was offered to children from this age group with high-risk conditions (at least 8% of the total population in this group).^14,15^ Different data sources and designs were used to compare the frequency of severe RSV infections (hospitalizations and ED visits) before (period varied according to data source) and after the start of the immunization program (season 2024-2025). Nirsevimab administration began on October 1^st^, 2024 (epiweek 40) for infants at increased risk of severe disease, then was offered to all eligible infants starting on November 4^th^, 2024 (epiweek 45). Although available in community pharmacies, the RSV maternal vaccine was not publicly funded, and uptake remained very low (< 3%); thus, children born to vaccinated mothers were not excluded from the analysis.^16^

### Data sources

Information on ED visits was obtained from Québec’s *Système d’information et de gestion des urgences* (SIGDU, an administrative information system). Hospitalizations were measured using three distinct data sources: 1) HospiVir (sentinel surveillance network in acute-care hospitals); 2) SIGDU (patients admitted at the end of their ED visit); 3) preliminary reports from the province’s acute-care admissions’ administrative database, MED-ECHO (*Maintenance et exploitation des données pour l’étude de la clientèle hospitalière*). SIGDU and MED-ECHO data were linked to RSV-positive tests from the provincial laboratory database. Data on RSV tests became available in the Fall of 2022. Demographic data (i.e. population per year and per age group) were obtained using the *Fichier d’identification des personnes assurées* de la *Régie de l’assurance-maladie du Québec*; in this data source, age is determined on October 1^st^ of each year.

Six hospitals admitting persons <18 years of age (including two pediatric tertiary hospitals) participate in HospiVir, covering around 30% of the province’s pediatric hospitalizations for ARI. Patients hospitalized for an ARI were eligible if they were admitted for at least 24 hours with one of the following symptoms: fever, cough (or worsening of a pre-existing cough), breathing difficulties (or worsening of breathing difficulties), recent onset of extreme fatigue or deterioration of general condition. All eligible cases were systematically tested with a multiplex PCR laboratory assay that included RSV testing. More details are available elsewhere. ^2,17^

### Variables

#### Intervention

The evaluated intervention was the 2024-2025 nirsevimab immunization campaign. Pre-intervention periods were defined according to data availability from different sources (Supplementary figure A). This decision to take as many pre-intervention years as possible aimed to obtain the best representation of possible seasonal variations. The intervention was implemented during the Fall of 2024, so that the 2024-2025 season was considered the post-intervention period. RSV seasons were defined by epiweeks when at least 5% of all provincial RSV tests were positive (Figure 1).

**Figure 1.**
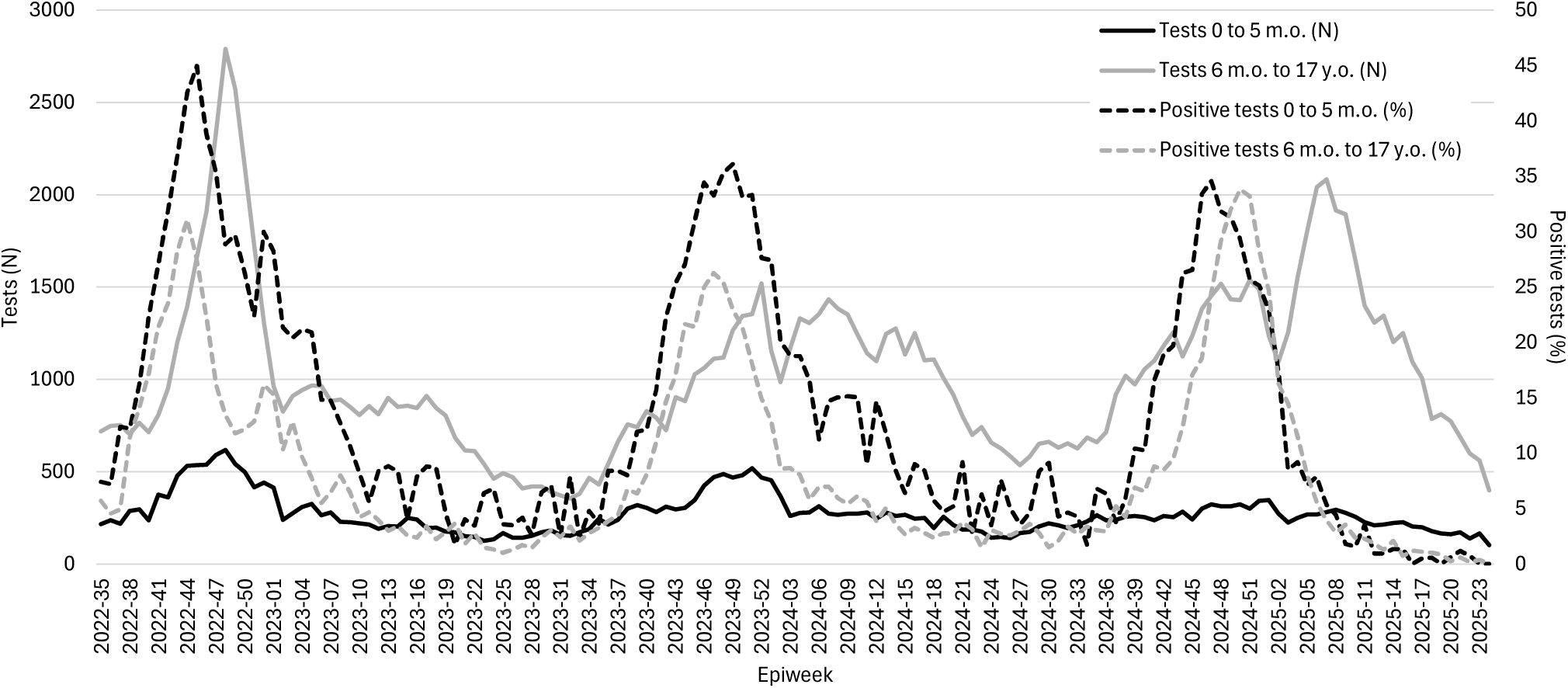
Weekly number of RSV tests and proportion of positive tests, by age group, Québec, 2022-2025.

#### RSV-confirmed and RSV-associated hospitalizations

RSV-positive hospitalizations identified in HospiVir were considered “RSV-confirmed hospitalizations”. For the validity of comparisons, only the 2023-2024 season was included in the pre-intervention period for RSV-confirmed hospitalizations, since the last hospital that joined the network, a pediatric tertiary hospital, was in 2023-2024. Demographic and clinical information were collected by reviewing the medical files after the patients’ discharge.

MED-ECHO hospitalizations that were linked to a positive RSV test 7 days prior to 3 days after admission were considered “RSV-associated hospitalizations”. This real-time administrative database includes information on age at admission, sex and duration of stay, while diagnostic codes to confirm the cause of admission are not available. Both the 2022-2023 and 2023-2024 seasons were included in the pre-intervention period.

#### ED visits for acute bronchiolitis and RSV-associated ED visits

SIGDU data include a single diagnostic code per ED visit, as well as information on age, sex and complications such as hospital admission or death. “ED visits for acute bronchiolitis” were defined as visits with a diagnostic code of acute bronchiolitis (ICD-10 code J21.9). Pre-intervention period covered years 2018-2019 to 2023-2024, excluding 2020-2021 and 2021-2022, because the COVID-19 pandemic resulted in two atypical RSV seasons. “RSV-associated ED visits” had a diagnostic code related to ARI,^18^ combined with an RSV-positive test 7 days before to 3 days after the visit. The 2022-2023 and 2023-2024 seasons were included in the pre-intervention period.

### Statistical analyses

Statistical analyses were conducted with OpenEpi 3.01, SAS 9.4 (SAS Institute, USA) and R (version 2024.12.1+563; R Foundation for Statistical Computing).

### Descriptive analysis of the study population

Demographic and clinical characteristics of included children before and after the introduction of nirsevimab were summarized. Continuous variables were reported as mean, median, first and third quartiles (Q1–Q3), and minimum and maximum values. Differences between pre-and post-intervention periods for continuous variables were assessed using the Wilcoxon rank-sum test. Categorical variables were summarized as counts and percentages, and comparisons between periods were performed using Pearson’s Chi-squared test (α = 0.05).

### Measures of severe cases frequency

The numbers and incidence rates per 100,000 population of all outcomes were measured for each age group. For RSV-confirmed hospitalizations, overall population estimates were adjusted for the proportion of the province’s pediatric ARI hospitalizations covered by participating hospitals (30%). For RSV-associated ED visits and ED visits for acute bronchiolitis, the proportion of visits concluding with the death or admission of the patient was also computed. For each indicator and age group, the relative difference between the pre-and post-intervention periods was estimated.

### Impact of the immunization program

#### Diference-in-diferences method (RSV-associated hospitalizations and ED visits, and acute bronchiolitis ED visits)

The variation in incidence rates or proportions among infants 0-5 months old before and after the implementation of the immunization program was compared to the corresponding variation among the non-eligible 1-17-year-old group (comparison group). Difference-in-differences allows for adjustment for generalized temporal trends (such as a more severe season) affecting all age groups, thereby isolating the impact of the intervention in the targeted group.

For RSV-associated hospitalizations, RSV-associated ED visits, and acute bronchiolitis ED visits, Poisson regression was used to model incidence rates. Robust Poisson regression was used to model proportions of complications in ED visits while also accounting for hospital-level correlation using generalized estimating equations. Regression analyses included the pre-versus post-intervention period, the targeted versus comparison group and an interaction term between these two variables; this last term quantifies the adjusted relative impact of the nirsevimab campaign [(1 – rate ratio (RR)) x 100]. Expected numbers, rates or proportions were estimated by multiplying the observed number of cases among infants 0-5 months old during the 2024-2025 season by 1 / RR of the interaction term. In sensitivity analyses, infants 6-11 months old were combined with those 0-5 months old. In other sensitivity analyses, only season 2023-2024 was included in the pre-intervention period, similarly to the analysis of RSV-confirmed hospitalizations.

#### Observed versus expected rates based on the relative weight of the targeted age group pre-intervention (RSV-confirmed hospitalizations)

The expected rate of RSV-confirmed hospitalizations in 2024-2025 in children 0-5 months old was estimated from the ratio of the RSV-confirmed hospitalization rate in this group to that in the 1-17 years old group observed in 2023-2024 (pre-intervention). This method is not affected by variations in severity of RSV seasons because it is based on the ratio of the targeted group relative to non-targeted groups, rather than the magnitude of the burden in the previous season used in before-after comparisons. The relative reduction was expressed as a coefficient of variation. A 95% confidence interval (CI) was estimated by bootstrap with discounting (10,000 iterations), using the 2.5^th^ and 97.5^th^ percentiles as interval bounds. Sensitivity analyses included only hospitals that had participated since the start of the network; pre-intervention seasons for these analyses were 2012-2019 and 2022-2024.

### Ethical Considerations

The analysis used deidentified data and was part of the program evaluation mandated by the Québec Ministry of Health. This project was exempt from the requirement for ethics approval by the CHU de Québec Review Board (#2026-8268).

## Results

### Demographic and clinical characteristics

Table 1 presents the main demographic and clinical characteristics of severe RSV cases during the pre-and post-intervention periods. Depending on data source, between 40% and 46% of cases were female. Around 12% of RSV-confirmed hospitalizations were born prematurely, and between 15% and 19% had comorbidities. Across all data sources, the median age at admission or ED visit was consistently higher post-intervention. In contrast, the median length of hospital stay was the same pre-and post-intervention, i.e. 2 days for RSV-confirmed hospitalizations and 3 days for RSV-associated hospitalizations. The proportion of ED visits resulting in hospitalization or death decreased in the intervention group: from 32.0% to 27.4% of visits for acute bronchiolitis, and from 39.0% to 31.6% of RSV-associated visits.

**Table 1:**
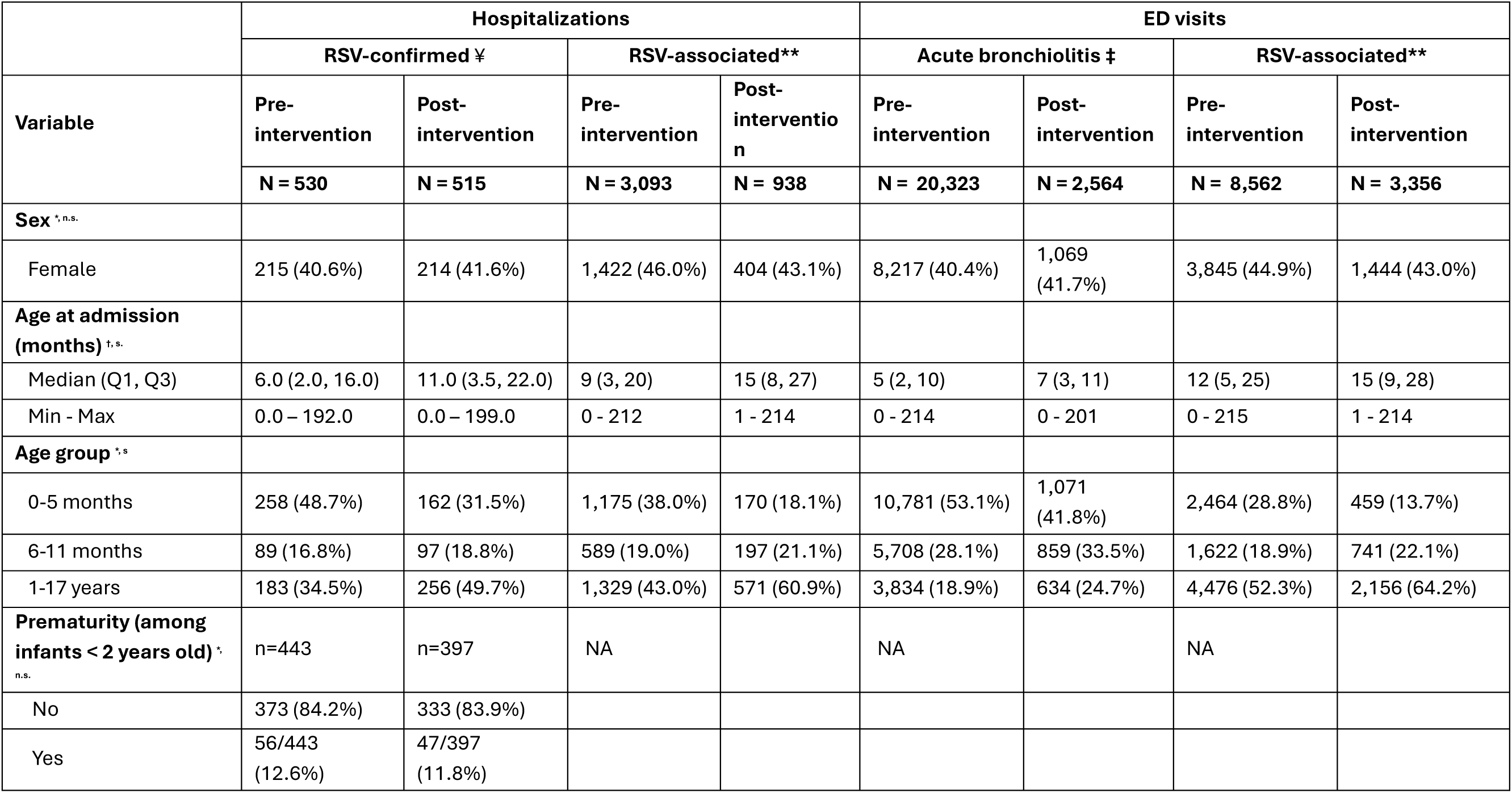
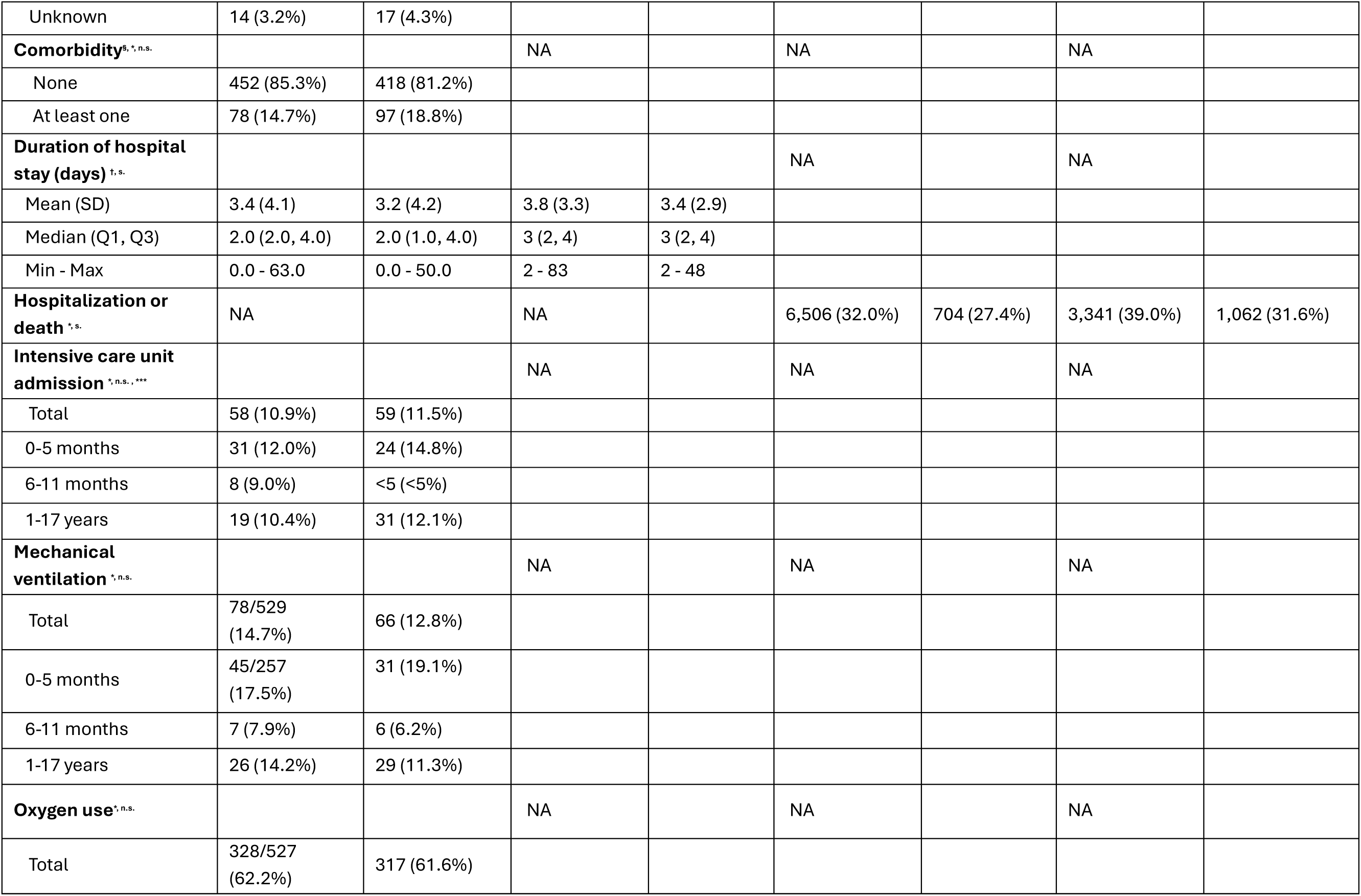
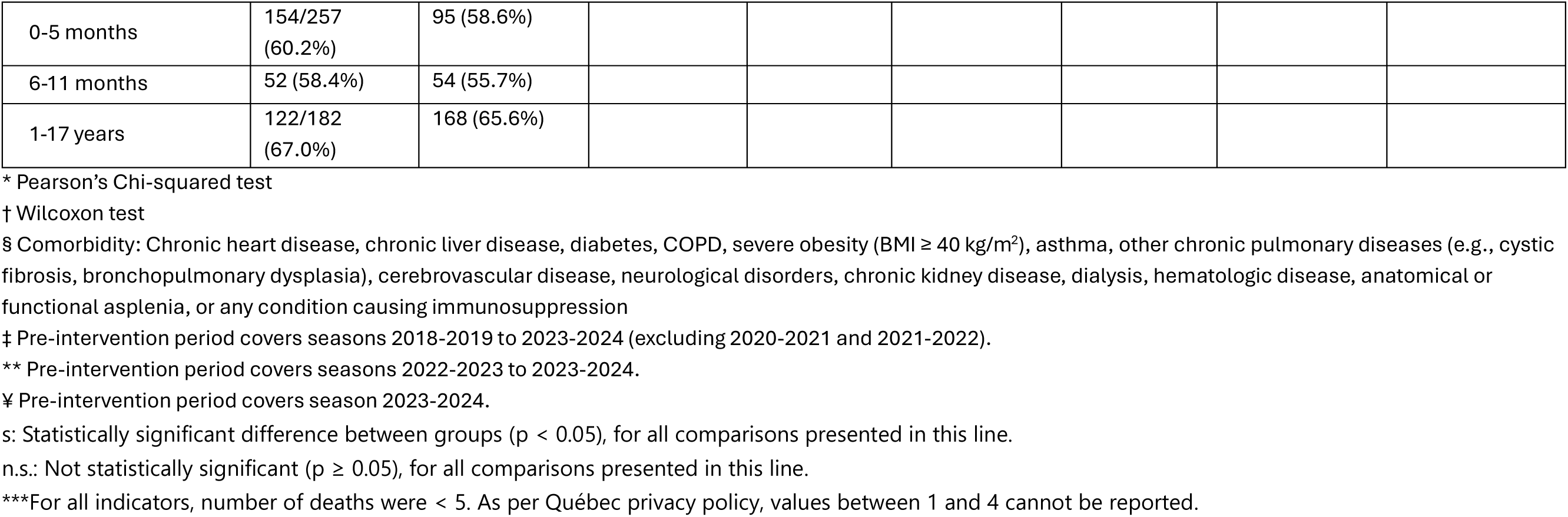
Comparison of demographic and clinical characteristics of RSV infections in children before and after the introduction of nirsevimab.

### Changes in RSV burden following nirsevimab introduction

Among infants 0–5 months old, the incidence rate of RSV-associated or confirmed hospitalizations was significantly lower post-intervention compared to the pre-intervention period (Table 2, Supplementary table A, Figure 2 and Supplementary figure B). A 37% reduction was observed in RSV-confirmed hospitalizations compared to 2023-2024 (RR: 0.63; 95% CI: 0.52–0.76), and a 71% decrease in RSV-associated hospitalizations compared to 2022-2024 (RR: 0.29; 95% CI: 0.25–0.34). In contrast, among children 1–17 years old, a 40% increase in RSV-confirmed hospitalizations (RR: 1.40; 95% CI: 1.16–1.69), and a 15% decrease in RSV-associated hospitalizations (RR=0.85; 95% CI: 0.77–0.94) were observed. For infants 6–11 months old, no significant changes occurred for RSV-confirmed hospitalizations, while RSV-associated hospitalization declined by 31% (RR=0.69; (95% CI: 0,58 – 0,81)).

**Figure 2.**
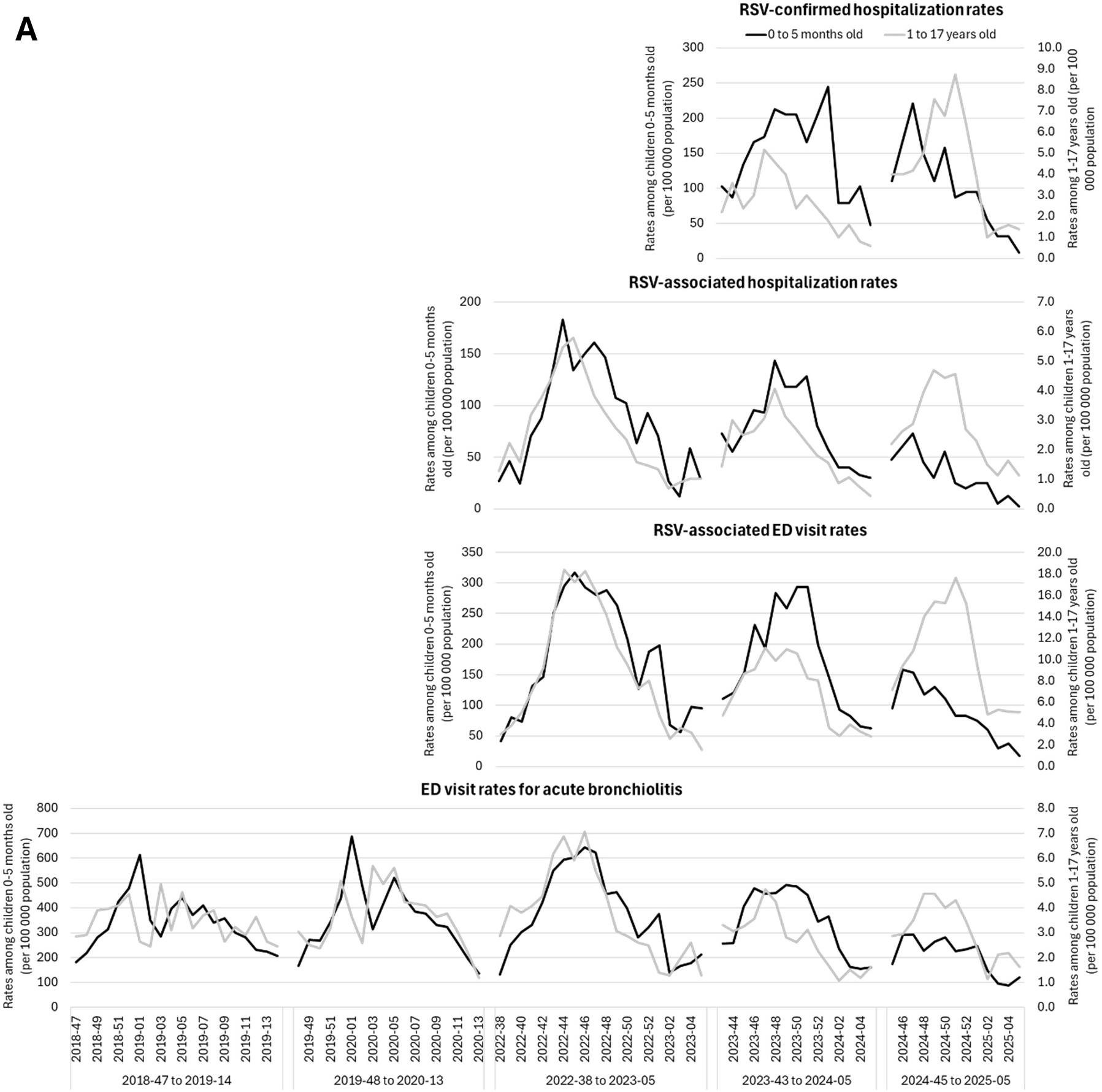
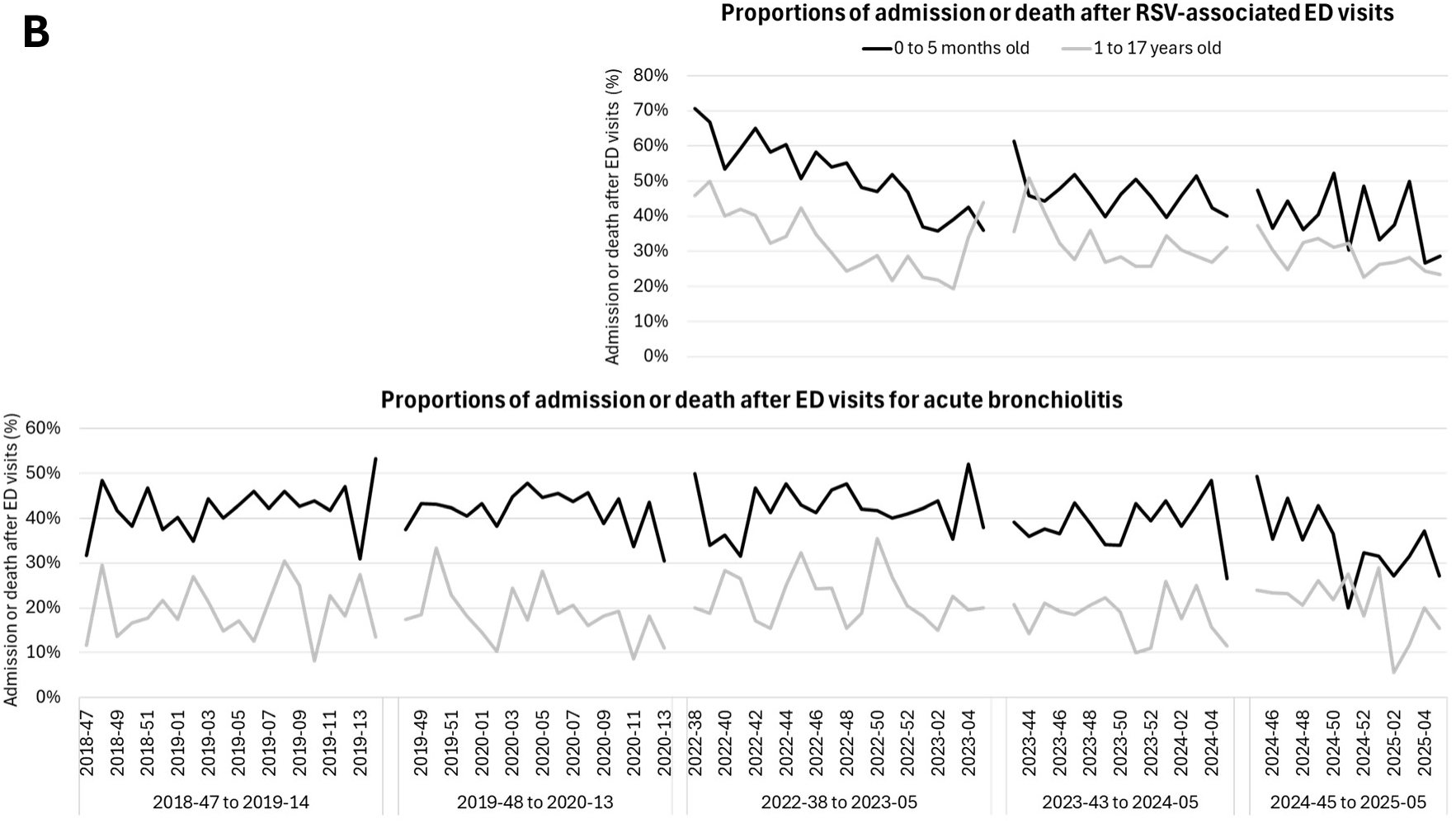
Weekly incidence rates of ED visits and of hospitalizations (panel A), and weekly proportions of hospitalization or death after ED visits (panel B), by age group and season, for ages 0–5 months and 1–17 years.

**Table 2.** Pre-and post-intervention diferences in the frequency of severe cases of RSV infections, by age group.

| Indicator | Rate or proportion in 0-5 months old<br>[95% CI] |  |  | Rate or proportion in 6-11 months<br>old [95% CI] |  |  | Rate or proportion in 1-17 years old<br>[95% CI] |  |  |
| --- | --- | --- | --- | --- | --- | --- | --- | --- | --- |
|  | Pre-intervention | Post-intervention | Post/<br>pre<br>ratio | Pre-intervention | Post-intervention | Post/<br>pre ratio | Pre-intervention | Post-intervention | Post/<br>pre ratio |
| RSV-confirmed hospitalization (rate per 100,000) <sup>¶</sup> | 2032.0<br>[1793.7;<br>2292.6] | 1275.9<br>[1088.0;<br>1486.6] | 0.63<br>[0.52;<br>0.76] | 772.0<br>[620.4;<br>949.1] | 841.4<br>[682.8;<br>1025.4] | 1.09<br>[0.82;<br>1.45] | 36.4 [31.3;<br>42.0] | 50.9 [44.8;<br>57.5] | 1.40<br>[1.16;<br>1.69] |
| RSV-associated hospitalization (rate per 100,000) <sup>§</sup> | 1453.7<br>[1370.6 ;<br>1536.8] | 426.8 [362.6 ;<br>490.9] | 0.29<br>[0.25 ;<br>0.34] | 743.3<br>[683.3 ;<br>803.4] | 509.3<br>[438.2 ;<br>580.4] | 0.69<br>[0.58 ;<br>0.81] | 41.9 [39.7 ;<br>44.2] | 35.7 [32.8 ;<br>38.6] | 0.85<br>[0.77 ;<br>0.94] |
| ED visits for acute bronchiolitis (rate per 100,000) <sup>‡</sup> | 6433.2<br>[6311.8 ;<br>6554.7] | 2688.7<br>[2527.6 ;<br>2849.7] | 0.42<br>[0.39 ;<br>0.44] | 3584.0<br>[3491.0 ;<br>3677.0] | 2220.8<br>[2072.3 ;<br>2369.3] | 0.62<br>[0.58 ;<br>0.66] | 61.9 [59.9 ;<br>63.8] | 39.6 [36.6 ;<br>42.7] | 0.64<br>[0.59 ;<br>0.69] |
| ED visits for acute bronchiolitis admitted or deceased (%) | 41.6 [40.7 ;<br>42.5] | 35.3 [32.5 ;<br>38.2] | 0.85<br>[0.78 ;<br>0.92] | 21.8 [20.8 ;<br>22.9] | 21.8 [19.1 ;<br>24.7] | 1.00<br>[0.87 ;<br>1.14] | 20.2 [19.0 ;<br>21.5] | 21.9 [18.9 ;<br>25.3] | 1.08<br>[0.92 ;<br>1.27] |
| RSV-associated ED visits (rate per 100,000)<sup>§</sup> | 3048.4<br>[2928.0 ;<br>3168.8] | 1152.3<br>[1046.9 ;<br>1257.7] | 0.38<br>[0.34 ;<br>0.42] | 2047.0<br>[1947.4 ;<br>2146.6] | 1915.7<br>[1777.8 ;<br>2053.7] | 0.94<br>[0.86 ;<br>1.02] | 141.2<br>[137.0 ;<br>145.3] | 134.8<br>[129.1 ;<br>140.5] | 0.96<br>[0.91 ;<br>1.01] |
| RSV-associated ED visits admitted or deceased (%) | 50.0 [48.0 ;<br>52.0] | 40.5 [36.1 ;<br>45.1] | 0,81<br>[0,72 ;<br>0,91] | 40.4 [38.0 ;<br>42.8] | 32.7 [29.4 ;<br>36.1] | 0,81<br>[0,72 ;<br>0,91] | 32.5 [31.1 ;<br>33.9] | 29.4 [27.5 ;<br>31.4] | 0,91<br>[0,84 ;<br>0,98] |

‡ Pre-intervention period covers seasons 2018-2019 to 2023-2024 (excluding 2020-2021 and 2021-2022).
§ Pre-intervention period covers seasons 2022-2023 to 2023-2024.
¥ Pre-intervention period covers season 2023-2024.
95% CI: 0.95 confidence interval.
ED: emergency department.

Consistent with hospitalization trends, ED visits for acute bronchiolitis and RSV-associated ED visits were lower in the post-intervention period among infants 0–5 months old, with reductions of 58% (RR= 0.42; (95% CI: 0.39–0.44)) and 62% (RR= 0.38; 95% CI: 0.34–0.42), respectively (Table 2). ED visits for acute bronchiolitis decreased in older children albeit to a lesser extent, while RSV-associated ED visits remained stable in older age groups. Additionally, the proportion of ED visits resulting in hospitalization or death in 0-5 months old infants was statistically lower by 15% post-intervention (visits for acute bronchiolitis, RR=0.85; (95% CI: 0.78–0.92)) and 19% (RSV-associated visits, RR=0.81; (95% CI: 0.72–0.91)).

### Quantification of the impact of nirsevimab on severe RSV-related cases

Observed weekly RSV-confirmed hospitalization rates were similar to expected rates from epiweeks 35 through 46 (Figure 3). However, beginning at epiweek 46 — approximately one week after expanding nirsevimab eligibility — a notable decrease in observed versus expected rates emerged. Rates of severe cases were lower than expected across all indicators (Table 3). RSV-confirmed hospitalizations decreased by 59% (95% CI: 49-70) and RSV-associated hospitalizations, by 66% (95% CI: 58–71). ED visits for acute bronchiolitis also decreased by 35% (95% CI: 28–41), and RSV-associated ED visits, by 60% (95% CI: 56–65). The frequency of ED visits that ended with the patient’s admission or death also decreased, by 25% (95% CI: 1-43) for acute bronchiolitis and by 20% for RSV-associated visits, although this reduction was not statistically significant (95% CI:-20-47). In all sensitivity analyses, although the estimated magnitude of the relative impact varied depending on the severity of seasons included in the pre-intervention period, interpretation did not change (Supplementary table B).

**Figure 3.**
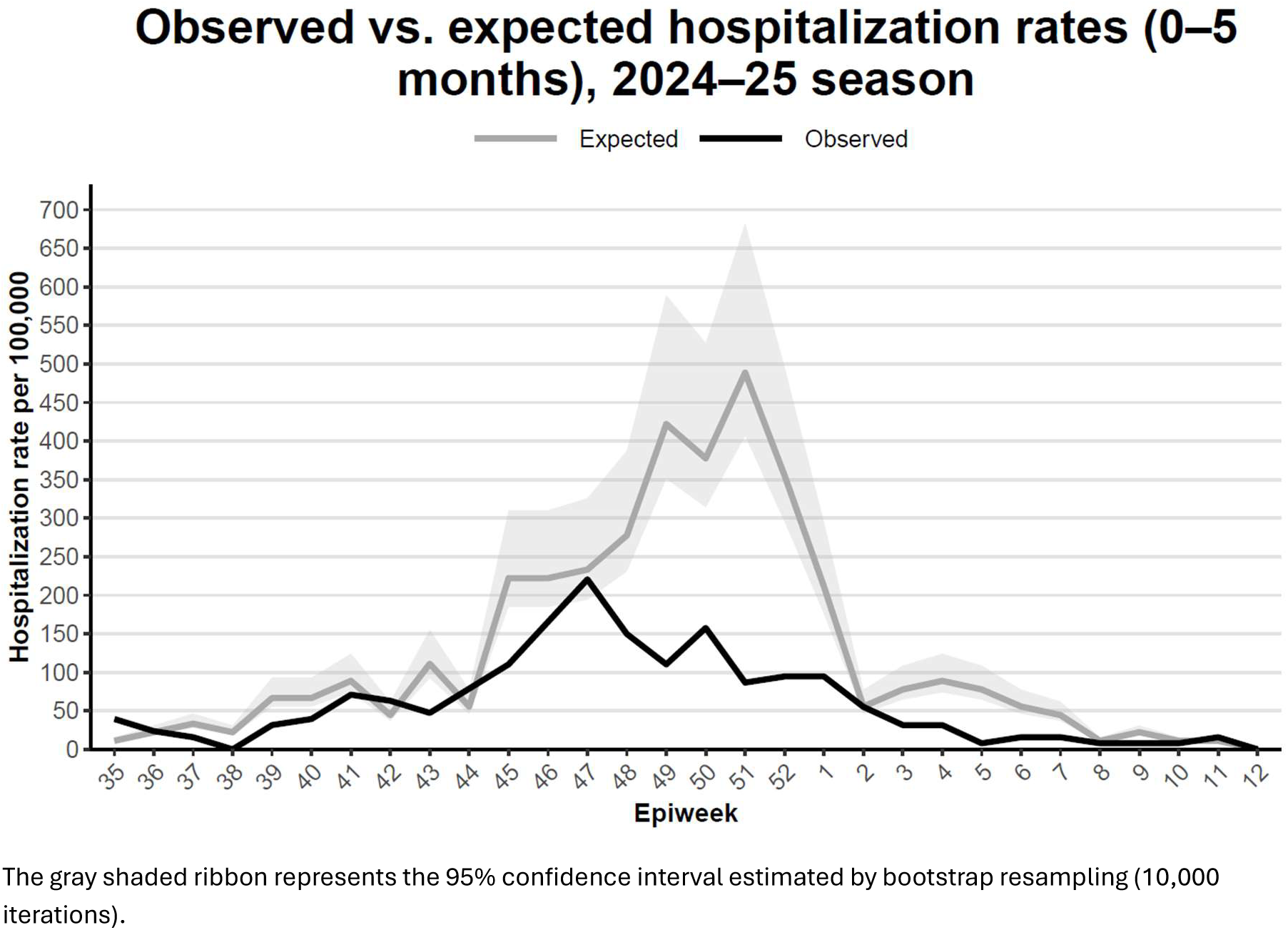
**Observed versus expected rate (per 100,000 population) of RSV-confirmed hospitalizations among children aged 0–5 months, post-intervention period, Hospivir, 2024-2025**

**Table 3.**
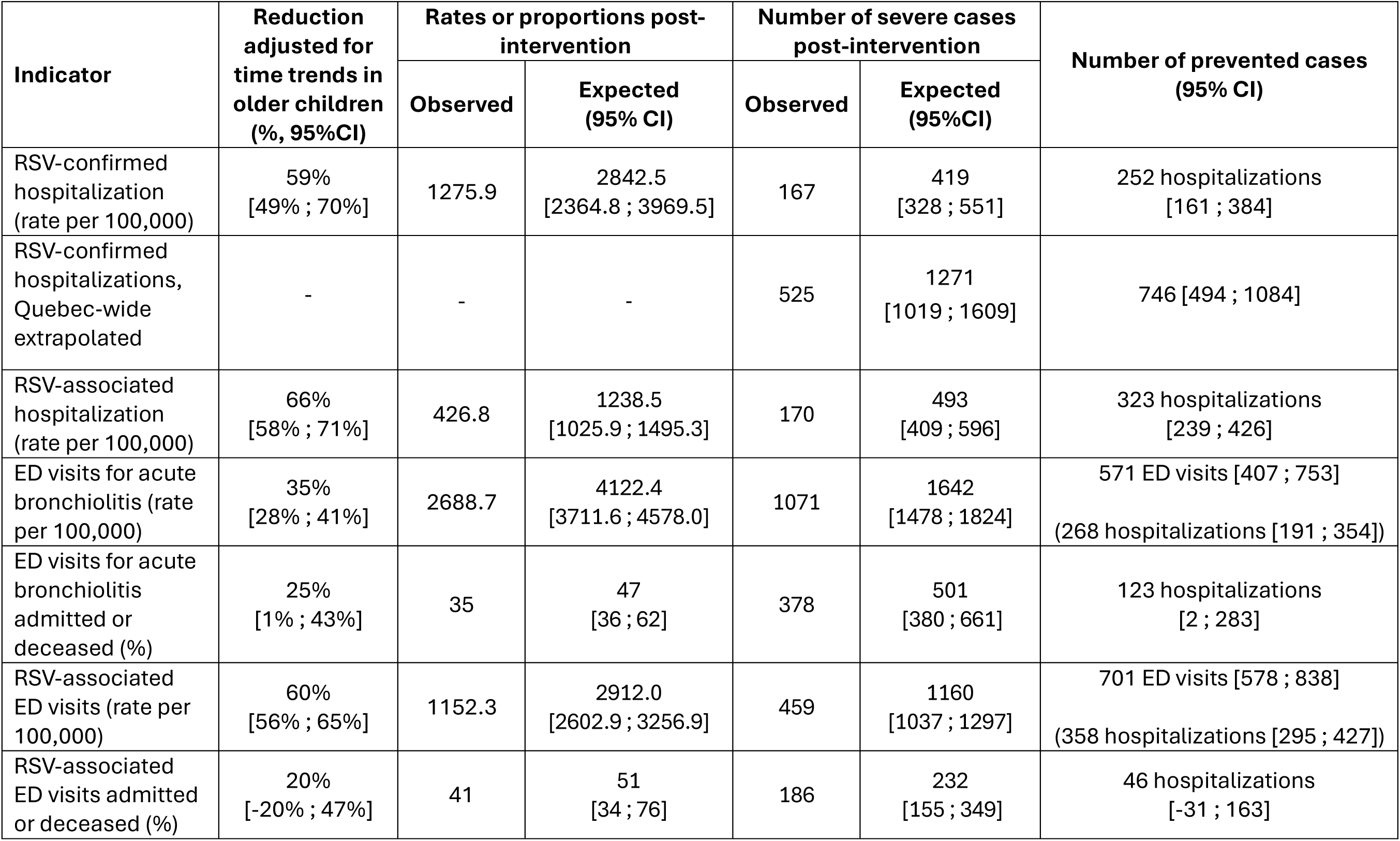
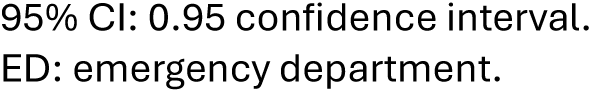
Reduction in observed severe cases in 0 to 5 months-old infants during the post-nirsevimab period, compared to expected numbers in the absence of an immunization program, after adjusting for trends in older children.

In terms of absolute numbers of prevented hospitalizations, active surveillance data provided higher estimates (Table 3; 746 RSV-confirmed hospitalizations) compared to administrative data (323 RSV-associated hospitalizations, 391 hospitalizations after an ED visit for acute bronchiolitis [i.e. 268 prevented visits that would have ended by hospitalization + an additional 123 admissions prevented through the reduced risk of hospitalization when an ED visit still occurred] and n = 404 [i.e. 358 + 46] hospitalizations after an RSV-associated ED visit).

## Discussion

In this study we applied two approaches to different data sources and definitions of severe RSV infections to measure the impact of Québec’s nirsevimab 2024-2025 immunization campaign. In the inaugural year of the immunization program, hospitalization rates among infants 0-5 months old were 59% to 66% lower than expected while adjusting for trends in older age groups without intervention. The risk of both ED visits for acute bronchiolitis and RSV-associated ED visits was also reduced by 35% and 60%, respectively, with an additional reduction in the risk of hospitalization for the remaining visits for acute bronchiolitis. Taken together, these temporal trends (adjusted for trends in comparison groups) suggest an early and significant reduction in RSV hospitalizations and ED visits attributable to the nirsevimab program’s implementation. While the maternal vaccine was also available for the first time in 2024-2025, given the very low uptake of RSV vaccination during pregnancy, it is unlikely that this contributed to the observed reduction in severe RSV cases. Overall, according to the different indicators of real-life evidence, several hundreds of hospitalizations would have been prevented with the 2024-2025 nirsevimab campaign among infants 0-5 months old, for a target population of around 40,000 babies. This estimation does not include the absolute numbers of hospitalizations prevented in 6-11-month-olds, who were eligible for nirsevimab at the beginning of the season but grew older and left the intervention group as defined in our study; therefore, the overall populational impact of the intervention, in absolute numbers, is underestimated in our study. Sensitivity analyses including this age group in the intervention group led to smaller relative impacts, as this group was not exposed to nirsevimab as much as 0-5 months old.

While the effect of immunization campaigns is frequently described in terms of effectiveness, overall populational impact reflects not only effectiveness, but also immunization coverage, herd immunity, access to care, clinician practices and ultimately better reflects of the public health benefits for the targeted population.^19–22^ This can lead to both different and valid estimates of the impact of nirsevimab in different populations. Indeed, a few studies have used designs similar to ours (covering a large population rather than a single hospital, targeting a similar population and adjusting for time trends in a comparison group not targeted by universal immunization). Estimated impacts vary, although most studies have reported larger impacts.^6–11^ In Galicia, with a 90% immunization coverage, a 90% reduction was observed in RSV-related lower respiratory tract infection hospitalizations among 0-5 months old children, during the first three months of the 2023–2024 RSV season compared to historical data, while disease frequency remained stable in older children.^23^ Another study observed an 87% reduction in RSV-positive hospitalization rates in Spanish 0-5 months old children, while a 29% increase was observed in 1 to 5 year-old children.^10^ In a third Spanish study based on a SARI sentinel network, RSV-related hospitalization rates were 74% lower in <1 year children after nirsevimab introduction in 2023-2024 compared to 2022-2023.^12^ In Ireland, RSV-related hospitalization rates of infants born during the 2024-2025 season were 75% less frequent than expected compared to the four previous RSV seasons, with an 83% coverage.^8^ In Luxembourg, an 84% coverage was associated with 69% less hospitalization numbers in babies under 6 months of age during the 2023–2024 season compared to the 2022–2023 season.^9^ Finally, in the US, where maternal vaccination was also offered, for a combined coverage against RSV of 66% in 0-7-months-olds, reductions in infant hospitalization rates during the 2024–2025 season compared to 2018–2020 ranged from 28% to 56%, while 7% to 64% increases were measured in 8-59-month-olds. ^11^ These studies all focused on hospitalizations, so our study provides a first estimate of the potential impact of nirsevimab on the burden of RSV-related ED visits.

In addition to program and coverage differences, measurement methodologies can affect nirsevimab impact estimations. With this study, we offer a comparison of results obtained with different data sources and varying lookback periods. Frequency measures, their relative variations as well as prevented cases are all relevant aspects to consider. HospiVir data (RSV-confirmed hospitalizations) is carefully validated by dedicated research nurses in all participating hospitals. However, because it covers only 30% of the provincial pediatric population, results have limited statistical power and projections are necessary to estimate the impact at the provincial level. On the contrary, population-based health administrative databases have strong statistical power but limited validation possibilities. For instance, when linking laboratory data with MED-ECHO or SIGDU data, the unique identifier is frequently missing for newborns, underestimating incidence rates. With MED-ECHO, successful linkages might also include RSV-positive patients admitted for other reasons, as no diagnosis is available, and it is unknown whether their propensity to be tested differs from that of other patients. Despite this last potential bias, RSV-associated hospitalization rates (MED-ECHO) are lower than RSV-confirmed rates (HospiVir), which suggests an overall underestimation of hospitalizations when RSV testing is not systematic. This underestimation should also apply to RSV-associated ED visits. However, we expect these biases to have been constant throughout the study period, allowing for a similar relative impact of nirsevimab with both MED-ECHO (66%) and HospiVir (59%), despite very different burden estimations and estimated numbers of prevented hospitalizations. Rates of ED visits for acute bronchiolitis showed a smaller relative decrease than other indicators, which is likely the result of being based solely on a diagnostic code that does not specifically identify RSV. Non-RSV episodes of acute bronchiolitis are also included in our indicator: these will not be prevented by nirsevimab, leading to an underestimation of the relative impact. Of note, all three estimations of prevented hospitalizations based on health administrative databases were similar, when considering confidence intervals. The use of a comparison group was also probably key in observing statistically similar nirsevimab impact across different lookback periods. In summary, active surveillance data probably provides better estimations of incidence rates and prevented hospitalizations, but its precision is limited; health administrative data can be used to measure nirsevimab’s relative impact, but it will underestimate incidence rates and prevented hospitalizations.

The real-life populational impact of immunization campaigns is best evaluated using ecological study designs, like ours.^24^ By design, ecological studies do not examine associations at the individual level (as opposed to vaccine effectiveness studies) and cannot lead to causal inference. In our study, an unknown factor might have lowered incidence rates of severe cases or RSV infections the same year as nirsevimab was introduced, specifically in infants 0-5 months old. This seems unlikely, as we used different indicators of severe cases, using different databases, and most results were consistent. The inclusion of a comparison population allowed for adjustment for potential temporal trends such as an increasing or decreasing propensity to test patients with acute respiratory infections or variations in season severity. As nirsevimab is not known to prevent RSV transmission, we assumed that no herd immunity effect has affected disease frequency in our comparison group.^25^ Analyses performed by week showed increasing program impact along with nirsevimab coverage.

## Conclusion

In Québec, the 2024-2025 nirsevimab immunization campaign was associated with a reduction by about two-thirds of RSV-related hospitalizations and RSV-associated ED visits among infants targeted by the program. ED visits for acute bronchiolitis were also reduced by a third. Quantification of RSV burden by using different sources, although differing in magnitude mostly due to underdetection, resulted in comparable estimations of the proportions of prevented hospitalizations. Nirsevimab has thus been key in preventing hundreds of severe RSV infections in Québec babies, an impact measured promptly after the most recent RSV season, thanks to access to real-time surveillance data.

## Data Availability

All data was obtained through governmental mandate and cannot be shared by the authors. Any requests should be addressed to the Ministère de la Santé et des Services sociaux du Québec.

