## Supplementary material for "Impact of the first nirsevimab immunization campaign on RSV-related emergency department visits and hospitalizations in Québec, Canada"

### Supplementary tables

**Table A. Number of severe RSV infection cases and population estimates* for rate calculation, pre- and post-intervention, by age group, Québec, 2018–2025**

| Indicator | 0-5 months old | | 6-11 months old | | 1-17 years old | |
|---|---|---|---|---|---|---|
| | Pre-intervention | Post-intervention | Pre-intervention | Post-intervention | Pre-intervention | Post-intervention |
| | (n / N) | (n / N) | (n / N) | (n / N) | (n / N) | (n / N) |
| RSV-confirmed hospitalization ¥ | 258 / 12697 | 162 / 12697 | 89 / 11529 | 97 / 11529 | 183 / 503161 | 256 / 503161 |
| RSV-associated hospitalization § | 1175 / 80829 | 170 / 39834 | 589 / 79238 | 197 / 38680 | 1329 / 3170933 | 571 / 1598997 |
| ED visits for acute bronchiolitis‡ | 10781 / 167583 | 1071 / 39834 | 5708 / 159264 | 859 / 38680 | 3834 / 6196017 | 634 / 1598997 |
| % admitted or deceased | 4485 / 10781 | 378 / 1071 | 1246 / 5708 | 187 / 859 | 775 / 3834 | 139 / 634 |
| RSV-associated ED visits§ | 2464 / 80829 | 459 / 39834 | 1622 / 79238 | 741 / 38680 | 4476 / 3170933 | 2156 / 1598997 |
| % admitted or deceased | 1232 / 2464 | 186 / 459 | 655 / 1622 | 242 / 741 | 1454 / 4476 | 634 / 2156 |

* Population estimates are based on a sample of the Québec population (see Methods)

† Rate per 100,000 population

‡ Pre-intervention period covers seasons 2018-2019 to 2023-2024 (excluding 2020-2021 and 2021-2022).

§ Pre-intervention period covers seasons 2022-2023 to 2023-2024.

¥ Pre-intervention period covers season 2023-2024.

**Table B. Sensitivity analyses in measuring the reduction in observed severe cases in 0 to 5 months-old infants during the post-nirsevimab period, compared to expected numbers in the absence of an immunization program, after adjusting for time trends in older children.**

| Modification | Indicator | Reduction adjusted for time trends in older children (%, 95%CI) | Rates or proportions post-intervention | | Number of severe cases post-intervention | | Number of prevented cases (95% CI) |
|---|---|---|---|---|---|---|---|
| | | | Observed | Expected (95% CI) | Observed | Expected (95%CI) | |
| Pre-intervention period includes 2012-2013 to 2018-2019, 2022-2023 and 2023-2024 seasons (only 4 hospitals) | RSV-confirmed hospitalization (rate per 100 000) | 69% [59% ; 75%] | - | - | 24 | 77 [58 ; 99] | 53 [34 ; 75] |
| Pre-intervention period includes only the 2023-2024 season | RSV-associated hospitalization (rate per 100,000) | 68% [61% ; 74%] | 426.8 | 1344.6 [1087.0 ; 1663.2] | 170 | 536 [433 ; 663] | 366 hospitalizations [263 ; 493] |
| | ED visits for acute bronchiolitis | 47% [40% ; 54%] | 2688.7 | 5117.4 [4483.3 ; 5841.1] | 1071 | 2038 [1786 ; 2327] | 967 ED visits [715 ; 1256] |

| | | | | | | | |
|---|---|---|---|---|---|---|---|
| | (rate per 100,000) | | | | | | (484 hospitlizations [358 ; 628]) |
| | ED visits for acute bronchiolitis admitted or deceased (%) | 29% [5% ; 47%] | 35 | 50 [37 ; 67] | 378 | 536 [399 ; 720] | 158 admissions [21 ; 342] |
| | RSV-associated ED visits (rate per 100,000) | 65% [61% ; 70%] | 1152.3 | 3336.1 [2938.0 ; 3789.2] | 459 | 1329 [1170 ; 1509] | 870 ED visits [711 ; 1050]<br><br>(418 hospitalizations [341 ; 504]) |
| | RSV-associated ED visits admitted or deceased (%) | 16% [-20% ; 41%] | 41 | 48 [34 ; 68] | 186 | 220 [155 ; 314] | 34 hospitalizations [-31 ; 128] |

95% CI: 0.95 confidence interval
ED: emergency department

### Supplementary figures

#### Figure A. Data sources and periods covered

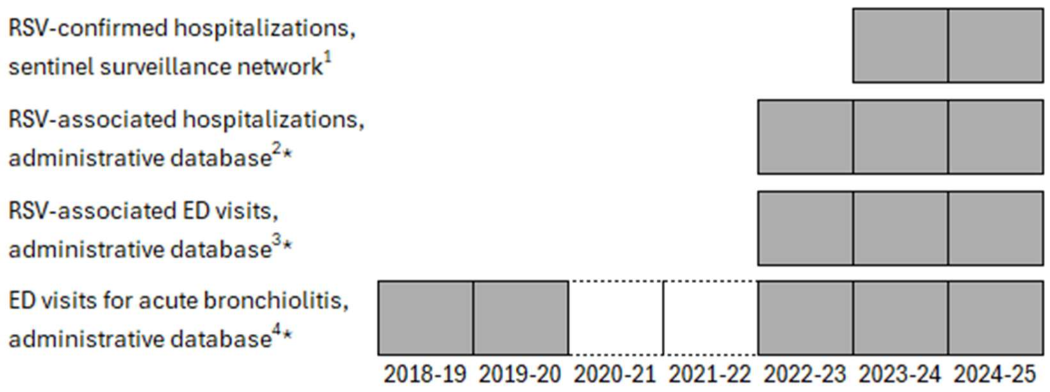


1. Patients hospitalized for acute respiratory infections (ARI), admitted for at least 24 hours with symptoms and who tested positive to RSV.
2. Patients hospitalized (regardless of diagnostic code, which were unavailable) and for whom a positive RSV test was obtained within 7 days prior and 3 days after admission.
3. Patients who presented at the ED for an ARI (based on diagnostic codes) and for whom a positive RSV test was obtained within 7 days prior and 3 days after the visit. Patients from this group who were then admitted or who died were also identified for a secondary indicator.
4. Patients who presented at the ED and received a diagnosis of acute bronchiolitis (ICD-10 code: J21.9). No laboratory test result was considered in this case definition. Patients from this group who were then admitted or who died were also identified for a secondary indicator. Due to the COVID-19 pandemic and associated public health measures, seasons 2020-21 and 2021-22 were atypical RSV seasons and were thus excluded from the study.

*For these indicators, cases were only counted for weeks when at least 5% of all RSV tests were positive epiweeks 2018-47 to 2019-14, 2019-48 to 2020-13, 2022-38 to 2023-05, 2023-43 to 2024-05 and 2024-45 to 2025-05.

**Figure B. RSV-confirmed hospitalization rates per 100,000 population, by age group and period (pre-intervention: 2023–2024; post-intervention: 2024-2025)**

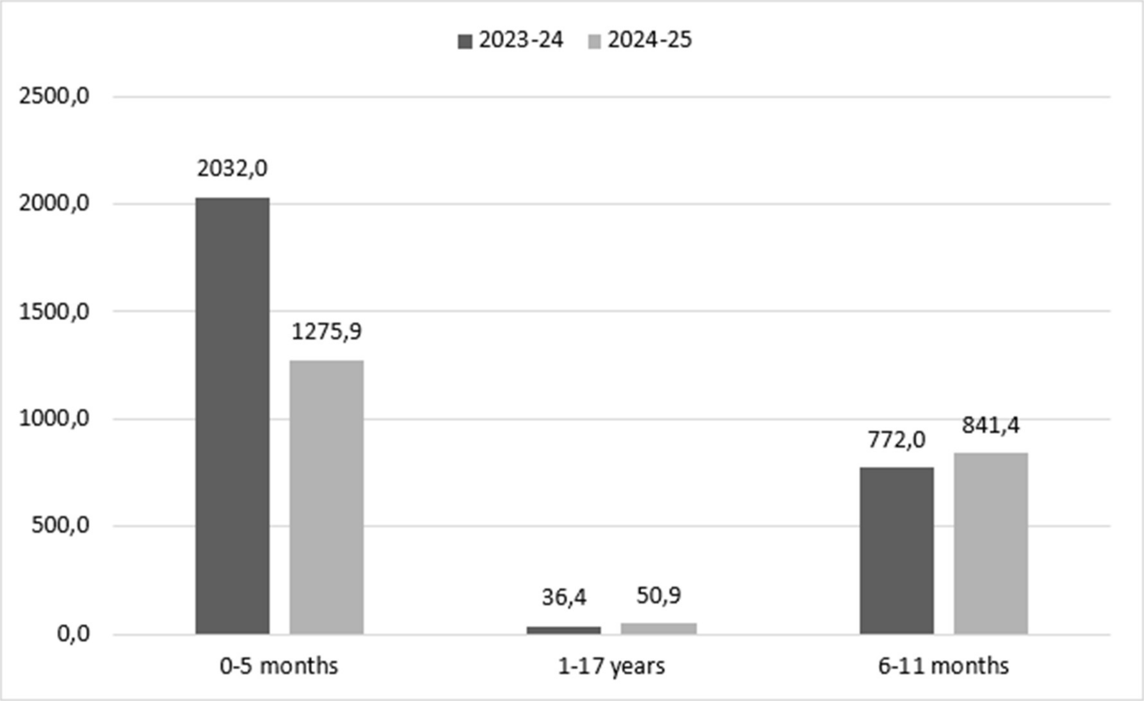
